# Establishing a national linked database for fetal alcohol spectrum disorder (FASD) in the UK: multi-method public and professional involvement to determine acceptability and feasibility

**DOI:** 10.1101/2024.03.26.24304887

**Authors:** Sarah K Harding, Beverley Samways, Amy Dillon, Sandra Butcher, Andy Boyd, Raja Mukherjee, Penny A. Cook, Cheryl McQuire

## Abstract

**Objective:** to conduct public and professional involvement work to establish stakeholder views on the feasibility, acceptability, key purposes, and design of a national linked longitudinal research database for fetal alcohol spectrum disorder (FASD) in the UK.

**Methods:** Following stakeholder-mapping, we identified contributors through collaborator networks and online searches. We consulted with stakeholders using online workshops (one for adults with FASD [and their supporters] N=5; one for caregivers of people with FASD N=7), 1:1/small-team video calls/email communication twith clinicians, policymakers, data-governance experts, third-sector representatives, and researchers [N=35]), and one hybrid clinical workshop (N=17). Discussions covered data availability, benefits, challenges, and design preferences for a national pseudonymised linked database for FASD. We derived key themes from the notes and recordings collected across all involvement activities.

**Results:** Our tailored, multi-method approach generated high levels of stakeholder engagement. Stakeholders expressed strong support for a pseudonymised national linked database for FASD. Key anticipated benefits were the potential for: increased awareness and understanding of FASD, leading to better support; new insights into clinical profiles, leading to greater diagnostic efficiency; facilitating international collaboration; and increased knowledge of the long-term impacts of FASD on health, social care, education, economic and criminal justice outcomes. Policymakers noted clear alignment with contemporary FASD and digital transformation priorities. Given the rich data infrastructure established in the UK, stakeholders expressed that a national linked FASD database could be world-leading. Common stakeholder concerns were around privacy and data-sharing and the importance of retaining space for clinical judgement alongside insights gained from quantitative analyses.

**Conclusions:** Multi-method and multidisciplinary public and professional involvement activities demonstrated the feasibility and acceptability of establishing a national linked database for FASD in the UK. Perceived benefits and challenges varied by stakeholder group, demonstrating that flexible, diverse, embedded stakeholder collaboration will be essential as we establish this database.

## INTRODUCTION

### Background

Fetal alcohol spectrum disorder (FASD) is one of the leading non-genetic causes of developmental disability worldwide [1]. It is thought to be particularly common in the UK, affecting an estimated 1.8 – 3.6% of children in the general population [2, 3]. Caused by prenatal alcohol exposure, FASD is associated with neurodevelopmental impairments, poor physical and mental health, substance misuse, and social problems across the lifespan [4–7]. Early diagnosis and support can significantly improve outcomes for those affected, as well as incurring significant cost savings for society [5, 6].

The full continuum of FASD consists of the subtypes ‘FASD with sentinel facial features’ (also known as Fetal Alcohol Syndrome [FAS]) and ‘FASD without sentinel facial features’[8].

Individuals who have FASD with sentinel facial features represent a minority of cases (approximately 10%) and present with small palpebral fissure length, smooth philtrum, and thin upper lip. Since most individuals with FASD do not have these recognisable facial features, FASD has been described as a mostly ‘invisible’ or ‘hidden’ disability [3, 8–10].

The invisibility analogy is consistent also with the lack of accessible data on FASD in the UK. This inaccessibility presents a significant barrier when attempting to achieve important FASD research, policy and healthcare goals [11, 12]. There are several reasons why UK data on FASD are currently lacking. First, standardised diagnostic codes for the full FASD continuum (FASD with and without sentinel facial features) are not readily available or used by clinicians. For example, all healthcare providers in England are required to use the Systematized Nomenclature of Medicine Clinical Terms (SNOMED CT) for capturing clinical information [13]. Although SNOMED CT does include a code for the full spectrum of FASD, only a code for fetal alcohol syndrome (i.e. FASD with sentinel facial features) is available for use in one of the main electronic clinical systems used in UK care settings - EMIS Web (EMIS Group plc©). Since approximately 90% of individuals with FASD do not present with sentinel facial features [3, 14]. FASD is significantly underreported. Furthermore, usage statistics show that the SNOMED CT codes that are available for FASD and prenatal alcohol exposure are not commonly used in practice [15]. Research investigating the reporting of FASD using Hospital Episodes Statistics (based on ICD-11 coding [16]) has similarly found high levels of underreporting as FASD is not easily detectable at birth, is not often a primary reason for hospitalisation, and under-recognition of FASD means that it is less likely to be coded as a co-existing condition [17, 18]. In addition, many clinicians report a lack of knowledge and confidence in making a FASD diagnosis and information on prenatal alcohol exposure is often missing or inaccurate, further complicating diagnosis. Perceived stigma, commonalities and comorbidities with other neurodevelopmental disorders, and the lack of a clearly defined care pathway have been cited as additional reasons for underdiagnosis and misdiagnosis of FASD [10, 13, 19–22].

There is a potential solution to addressing this critical FASD ‘data gap’. In 2019 the Scottish Intercollegiate Guidance Network (SIGN) published landmark guidance on children and young people with prenatal alcohol exposure [8]. Since this publication, there has been a swathe of complimentary policy and guidance publications in the UK. Many are now in effect nationally, including a National Institute for Health and Care Excellence (NICE) Quality Standard for FASD [11] and Department of Health and Social Care (DHSC) Health Needs Assessment for FASD [14], among others [23, 24]. These publications represent a unified call for a step-change in provision for FASD, including increased prevention, awareness, understanding, diagnosis, and support [8, 11, 14, 23, 24]. In parallel, there is a drive for digital transformation across the UK to improve health and social care services. This includes the Government’s ‘Data Saves Lives’ policy, which envisages a landscape of regional ‘secure data environments’ linking together electronic health and care records to enable analyses for the public good [25–31]. Data on FASD are currently being collected in some NHS and private clinical settings across the UK and publication of the aforementioned FASD guidance is anticipated to lead to a significant increase in diagnostic provision [23], with several new FASD clinics due to open in the near term. This could make a register utilising local NHS data feasible for the first time.

International exemplars are available. FASD databases in Australia, Canada and the USA have catalysed advances in diagnosis, treatment, prevention and support [32–37]. We aspire to achieve the same in the UK. Moving beyond international exemplars, established UK data linkage infrastructure offers potential for an electronic longitudinal linked database for FASD. This would be the first of its kind in the world. As highlighted by Health Data Research UK (HDR UK) “The UK is in a unique position to realise the potential of [routinely collected] health data, thanks to the NHS and its cradle-to-grave records for a population of over 65 million people” [38]. In this context, we have a crucial and timely opportunity to establish a world-leading data infrastructure for FASD in the UK that is efficient and standardised, can be linked to routine data, and which would be pivotal in informing, monitoring progress against, and achieving, (inter)national policy and research objectives [10, 11, 23, 39].

### Aim and scope

Stakeholder support is crucial for the success of large-scale data research projects, such as this [40]. One way to achieve this is through early, sustained, and meaningful patient and public involvement (PPI) [41, 42]. In this paper, we follow the National Institute for Health and Care Research definition of PPI, as follows: “*research being carried out ‘with’ or ‘by’ members of the public rather than ‘to’, ‘about’ or ‘for’ them”* [43]. The work that we present in this paper represents the earliest stages of the research cycle model, namely: prioritising/shaping research, research design and grant development [43].

The primary aim of this PPI project was to work with relevant public (adults with FASD and their caregivers) and professional (clinicians, data experts, third sector representatives, researchers and policymaking) stakeholders to establish the feasibility, acceptability, key purposes, and design of the first pseudonymised linked national longitudinal research database for FASD in the UK.

## METHODS

### Approach

Our PPI activities were designed with consideration of the UK Standards for Public Involvement in Research (UKSPI) [44] and reported according to the GRIPP2 (Guidance for Reporting Involvement of Patients and the Public) checklist [45]. The project team included individuals from academic, clinical, third sector and data governance sectors.

### Approvals

Ethical approval and the completion of consent forms was not required as this project was classed as PPI. All stakeholders were informed of the purpose of the PPI activities and were told that their participation was voluntary. Workshop contributors were informed that their anonymised data may be used in project outputs, and that they had the right to opt out of being included in recordings and outputs.

### Stakeholder identification and engagement

We conducted stakeholder mapping to identify key groups and contacts for PPI, following methods outlined in [46]. Contacts were identified through the project team’s networks and online searches. Following USKPI principles, we sought to provide flexible and inclusive opportunities for PPI. Below we describe our PPI approach for each of our stakeholder groups. Figure 1 provides a visual summary of our involvement activities over time.

**Figure 1:**
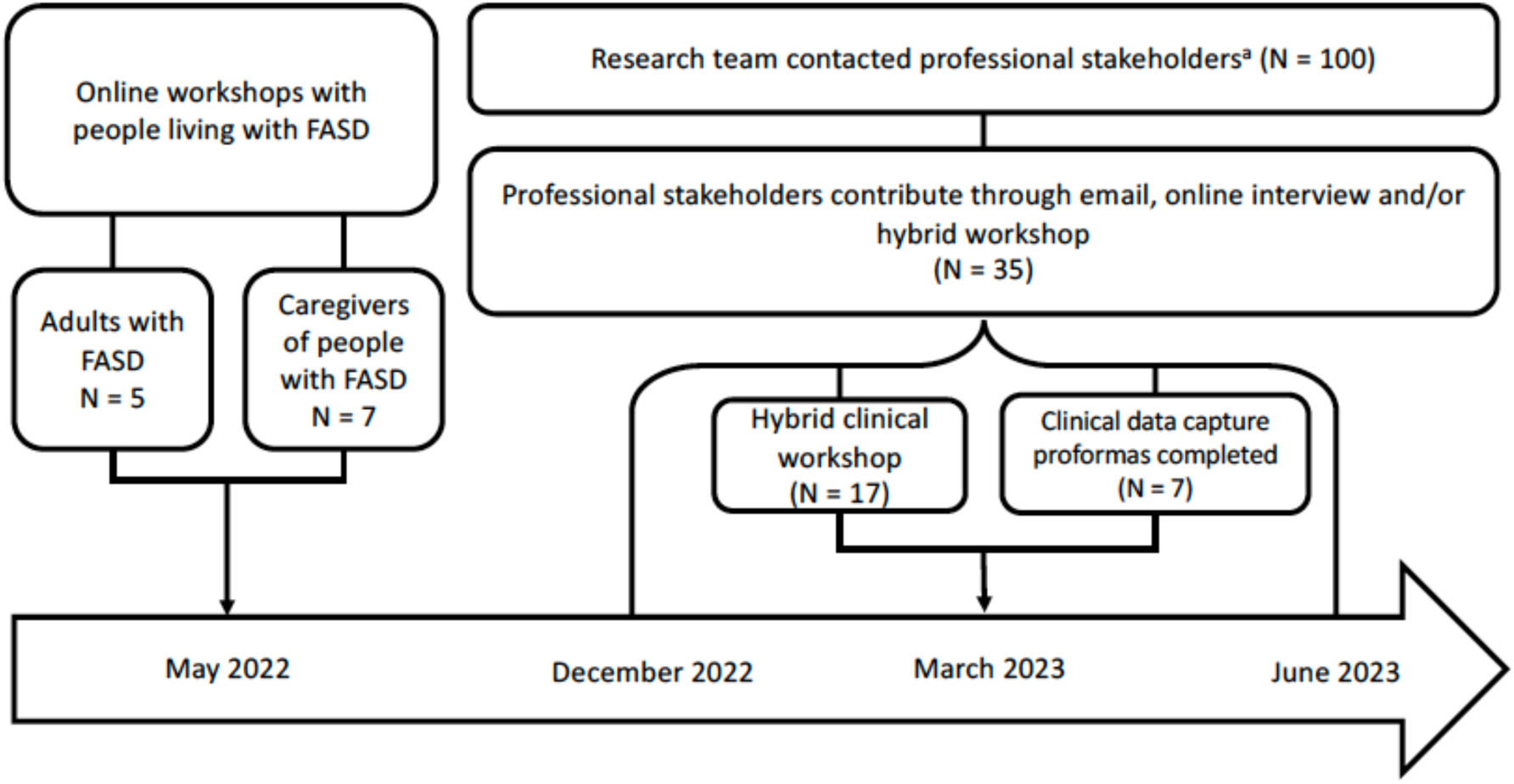
Summary timeline of all public involvement activities related to the development of a National Database for Fetal Alcohol Spectrum Disorder (FASD) in the UK. ^a^ Professional stakeholders included third sector, clinical, academic, data providers/specialists, and policy representatives.

### ‘Lived experience’ stakeholders: Adults with FASD and their caregivers

The format, materials, identification of public contributors, and delivery of our PPI activities with ‘lived experience’ stakeholders (adults with FASD and their caregivers) were designed and delivered in collaboration with the National Organisation for FASD and an adult with FASD and their birth mother. Online workshops were deemed to be the most appropriate format for involving stakeholders with lived experience. Contact with adults with FASD and their caregivers was supported by the National Organisation for FASD, who assisted with the design and distribution of easily accessible visual information sheets and flyers promoting the PPI opportunity through their email distribution lists and social media channels.

Members of the project team also circulated details of the opportunity through social media. Stakeholders with lived experience who were interested in taking part contacted the study team via email or telephone to confirm participation and were given easy access information about the proposed database and the opportunity to ask further questions before agreeing to take part. Adults with FASD were invited to have a caregiver with them for support during the online workshop if they wished. All contributors with lived experience were reimbursed for their contribution following the NIHR payment guidance for researchers and professionals [47].

We carried out two one-hour online workshops using videoconferencing software in May 2022, one for adults with FASD and their supporters (N=5 adults with FASD), and one for caregivers of people with FASD (N=7). Workshops were facilitated by CM and SB and sought to explore the views and experiences of people with FASD and their caregivers with regards to: a) data sharing and consent for research, particularly in relation to views on a national FASD database and use of their personal data; b) research priorities; c) procedures for the study; d) whether they would like to be contacted to be involved in the study in the future as a contributor or PPI/steering group member; and e) any other aspects that contributors deemed relevant.

### Professional stakeholders: third sector, clinical, academic, data providers/specialists, and policy representatives

Professional stakeholders were contacted by members of the project team via email or webform, provided with information about the PPI purpose and activities, and offered the opportunity to be involved through email, 1:1/group videocalls and/or a four-hour hybrid workshop. Videocall and email discussions were tailored to maximise relevance for each stakeholder and covered topics including data availability, consent and governance issues, and data storage/linkage opportunities. Informal notes were made of these discussions by members of the project team (CM, BS and AD). Between December 2022 and June 2023, we contacted 100 professional stakeholders, of whom 35 engaged in videocalls and/or email communication. We hosted the hybrid PPI workshop at the University of Salford in March 2023. This workshop took place the day before the ‘FASD in the UK’ conference (held at the same location) to maximise the potential for in-person workshop attendance. We provided in-person attendees with refreshments and travel reimbursement. The workshop was designed primarily for clinicians and included opportunities for: networking; an introduction to our proposal for a pseudonymised national linked longitudinal database for FASD; presentations from the Director of the UK Longitudinal Linkage collaboration (AB) and lead for the Canadian National Database for FASD (Professor Jocelynn Cook); breakout and full-group discussions on data sharing and governance; clinical data capture; presentation of a draft data pipeline for a national FASD database; and discussion of perceived benefits and challenges of establishing and using a national database for FASD in the UK. Seventeen stakeholders attended the workshop (ten in-person and seven online attendees consisting of 16 clinical and one third-sector representative). To assess the availability and format of clinical data on FASD, clinical stakeholders were also invited to complete a proforma, based on the Canadian National Database for FASD data capture form (template shown in Supplementary Appendix 1). The proforma included a list of measures relating to FASD including demographic information, FASD symptomology (including details of neurodevelopmental outcomes, growth and sentinel facial features), and assessment of parental alcohol exposure. For each measure, contributors were asked whether this type of data was currently being collected in clinic, whether they would consider collecting it in the future and if they would be willing to share the data (subject to robust governance).

### Synthesis

Two members of the project team (SH and CM) familiarised themselves with the notes and recordings collated for all PPI activities and produced summaries of the key themes discussed across stakeholder groups.

## RESULTS

Public and professional contributors’ discussions on the feasibility and acceptability of a pseudonymised national linked database for FASD were grouped according to the following three themes: i) perceived benefits, ii) perceived challenges, iii) recommendations for database design. These results are summarised in Table 1 and described further below.

**Table 1:** Summary of themes across all public and professional involvement activities.

| Theme | Sub-theme (if applicable) |
| --- | --- |
| **Benefits of establishing a national database for FASD in the UK**<br>*'advantages all around'* | • Advancing FASD research and policy<br>• Increased awareness, understanding, and support for FASD<br>• Informing clinical practice<br>• Supporting people living with FASD 'to be heard' |
| **Challenges of establishing a national database for FASD in the UK**<br>*'a mishmash of data'* | • Data sharing, governance, privacy considerations<br>• Establishing the most appropriate model of consent<br>• Existing data are inconsistent, unreliable, inaccessible<br>• Difficulties of reducing complex diagnostic data<br>• Limited in-clinic resources for data input |
| **Recommendations for database design**<br>*'numbers don't always tell the truth'* | • Needs to be manageable for busy clinicians/staff<br>• Should include confirmed and unconfirmed FASD cases<br>• Should enable input of qualitative, as well as standardised quantitative data<br>• Should include a 'full picture' of FASD |

Under the ‘data availability’ subheading at the end of this section, we describe the availability and format of FASD data, based on the data capture proformas completed by clinical contributors.

### Benefits of establishing a national database for FASD in the UK

Across all groups, public and professional contributors were supportive of a national database for FASD in the UK, speaking of ‘*advantages all around’.* Key anticipated benefits included advancing research and policy, informing clinical practice, increasing diagnostic efficiency, and the potentially ‘*endless possibilities*’ for prompting increased understanding, awareness, and support for FASD. In the excerpt below, a clinician recalls a phrase that they heard from people living with disabilities to express these ideas:

> *‘The message was “no data, no problem, no actions” …any attempt to bring data together allows us all to speak from something other than our opinions, because that doesn’t quite cut it… being stronger together would let us have a voice more importantly the individuals with that lived experience and exert their right to be heard through having data collected about them with them’ (Clinician P6, main workshop)*

In addition, people living with FASD described the burden of having to explain their condition to others and noting that:

> *‘[there is] so little proper joined up understanding of FASD’ (Caregiver of person with FASD, online focus group)*

> *‘trying to explain my FASD is not easy for someone who hasn’t got a clue, [it] would save me a lot of stress to explain it.’ (Adult with FASD)*

They suggested that a national database for FASD might help with this, shedding light on this often ‘hidden’ condition, and facilitating improved access to support.

> *‘I feel like I’ve been in the shadows for a while. If it’s going to help the situation, open doors, then maybe it’s best to share [the data]’ (Adult with FASD)*

> *‘My son has so much more difficult time than those with ADHD, autism, etc… why is FASD still not making a difference in what support [he gets]?’ (Caregiver of child with FASD)*

However, during the online workshop, it became clear that the relatively abstract concept of an anonymised database may be difficult to grasp for some adults with FASD. For example, some mistakenly thought that the database would be available to healthcare practitioners when they went for medical appointments and that this was how the database would help others to understand their condition. This demonstrated that further refinement of our materials and involvement activities with people with FASD will be needed to fully understand their views and preferences for a national database for FASD in the UK.

Academic contributors were enthusiastic about the potential for a national database for FASD to illuminate the long-term outcomes and costs associated with FASD, potential opportunities for intervention, and cost-benefit analyses of improved prevention, diagnosis, and support. Policy contributors noted clear alignment with contemporary FASD and data-transformation policies.

Clinical stakeholders discussed the potential benefits of a national FASD database for providing novel insights into clinical profiles, which in turn could lead to improvements in FASD diagnosis processes:

> *‘I can see how this database will be a potential way of structuring and really helping develop assessment more nationally’. (Clinician, P4, workshop breakout session)*

Stakeholders noted that these clinical insights could lead to significant gains in the efficiency, and therefore capacity for, FASD diagnosis, which is currently a time-consuming and complex process, and one which has an extensive waiting list. For example, if the data could help identify which brain domains are most commonly impaired, this could lead to a minimum data set for assessment, thus streamlining the process, as described in this exchange:

> *‘…some of these factors are really robust…the [Canadian] data really show those brain domains that are always, always, well 80% impaired…that’s pretty powerful’ (Researcher, P17, workshop)*

> *‘What [P17] just said about the [commonly affected brain] domains was so powerful…one of the questions that comes up constantly is “we can’t do multidisciplinary assessment”…if there are three [neurological] domains that come up constantly we could be able to make a diagnosis, particularly the ones that come up that are easy to [assess]…if that doesn’t come up then go to multidisciplinary assessment because we have to make a cost-effective model which actually works…these big datasets allow you to say “look this is what it’s actually showing”’ (Clinician, P9, main workshop)*

Taken together, these discussions highlighted the potential to support a tiered assessment model to enable people with FASD to achieve a robust diagnosis, more quickly and cost-effectively, with more ‘*complicated*’ cases being referred for further assessment. While this ‘*slim[med] down*’ model was broadly endorsed by clinical stakeholders, some also raised concerns that these data-driven insights could lead to an overly-simplistic approach for what can be considered a complex disorder. These views are described in more detail in the *challenges* section below.

### Challenges in developing a national database for FASD in the UK

While contributors were supportive of a national database for FASD overall, they also anticipated some challenges, explaining:

> *‘I think the reason we’re all thinking challenges is because we want to happen and therefore we’re trying to anticipate any hiccups on the way.’ (Clinician P5, main workshop)*

Clinical stakeholders expressed the importance of ensuring that any variables included on a future National FASD Database are not viewed as the only or most important factors to be considered in clinical assessment:

> *‘One of the challenges about [data standardisation] might be to…accidentally imply…that people can only assess with certain tools…Because this database can become a really powerful and effective tool we don’t want to just accidentally go down the road, that if it’s not on the database, it doesn’t matter.’ (Clinician P4, main workshop)*

Contributors from the clinical workshop noted that data recording processes were highly variable across settings, and that missing data were common, often because clinicians do not have the time and resources to collect and input the data. Data were recorded in paper and electronic format and those who use paper records expressed concerns about the cost and resources required to convert data to electronic records:

> *‘we have [two electronic] database[s]…and…paper folders…I’m the only person who’s [adding to the electronic records] so it’s not going very fast…Some things are added. Some things aren’t. It’s a weird mishmash…there’s no kind of real reason to what’s on there.’ (Clinician P7, breakout room 2)*

Establishing an appropriate model of consent was discussed as another key consideration. The majority of contributors expressed that it would be acceptable, and even preferable, to establish a model where relevant information on FASD cases from participating clinics was included in the database by default (i.e. an opt-out model) stating that they would:

> *‘feel fine about [data sharing] with reassurance of confidentiality’ (Caregiver of person with FASD)*

Furthermore, clinical, academic and data governance contributors expressed a strong preference for a default (opt-out) consent model on the basis of ‘public good’ and to ensure the sustainability of the database, given the practicalities of this particular population.

Clinicians from settings with existing FASD data explained that while they did have consent to contact some patients for future research studies, obtaining consent from these individuals would be time-consuming and potentially challenging as, for example, individuals with FASD are more likely to experience unstable living arrangements, and/or be involved with the care or criminal justice system. Academic contributors advised that a default (opt-out) model of consent would be important to minimise sampling bias and to ensure that participants who were ‘hard to reach’ and who, arguably, may be most in need would not be excluded.

While most contributors expressed a preference for a default consent model, some people living with FASD expressed reservations and indicated that they may prefer an opt-in model:

> *‘I’d just rather have someone ask. We’ve never had someone ask what we would prefer. To know it was used behind my back. Would like to give permission for research. Would bug me if they didn’t ask.’ (Adult with FASD)*

In terms of onward linkage, most adults with FASD and their caregivers suggested they would be comfortable with potential linkages from a national FASD database to educational, health, criminal justice and employment data. Contributors with data linkage expertise expressed the importance of ensuring that there was not a risk of reidentification, particularly as the number of cases of FASD in a national database may be small initially.

All contributors emphasised the need for robust data governance processes. Some clinicians from private settings stated that they would need additional support from data governance/legal experts when developing data sharing protocols, as they did not have a dedicated information governance team:

> *‘I want to share my data. I don’t have a problem, but the legal responsibilities that I have are really high…and because I’m not part of the NHS or a bigger culture it literally does stop with me, so I suppose that’s something I would need assistance with.’ (Clinician P10, main workshop)*

Together, these discussions show that while stakeholders were supportive of the database overall and deemed it to be in the public interest, ongoing engagement with those living with FASD in the design and communication of consent processes would be required to deliver this data resource in a transparent and accessible way. Additionally, clinicians highlighted resource constraints that may impact the feasibility of data collection and input. Stakeholders’ suggestions on how to address some of these challenges will be addressed further in the next section.

### Recommendations for the design of a national database for FASD in the UK

Public contributors discussed the design elements that they felt were important to consider when designing a national database for FASD in the UK. Key points were that data collection should be standardised so far as possible, but should also leave room for qualitative input; that contributing to the database needs to be manageable for clinic staff and may require further resources to support data input; that the database should to provide a ‘full picture’ of the lives of those with FASD, including strengths and difficulties; and that data should include both confirmed and unconfirmed FASD cases. These points are described further below.

First, clinicians noted that a degree of data standardisation and harmonisation would be valuable, stating:

> *‘you need to get consensus on what assessment will you use for what domains and have as much standardisation [as possible]’. (Clinician P6, main workshop)*.

Standardisation was also seen as important for facilitating comparison with international FASD datasets:

> *‘I think we can absolutely do international collaborations…the more data you have, the better we are’ (Clinician P17, main workshop)*

Data governance and clinical contributors also discussed the feasibility of sharing FASD data between different countries in the UK, for example between England and Scotland. Due to difficulties with ‘cross-border’ data sharing processes/infrastructure, contributors felt it was most feasible, in the first instance, to collate FASD data independently in England and Scotland, and then determine whether this approach can be adapted over time. It was agreed that a standardised data collection protocol would help to ensure that data are comparable if it is possible to merge English and Scottish FASD datasets in the future. This could include the use of a standardised data capture proforma (such as that provided in Supplementary Appendix 1).

Clinicians recommended that consent for the database should be flexible to allow for any future projects:

> *‘here the challenge… is prospective use of the data because things change over time and it’s good that that permission has some flexibility in it, which allows an important new data set which we haven’t thought of today’ (Clinician P9, main workshop)*

People with lived experience of FASD expressed that they wanted to ensure that any linked data would provide a full picture of the lives of those with FASD, including both strengths and difficulties. For example, caregivers of people with FASD highlighted that people with FASD may be ‘vulnerable’ to victimisation and noted that any linkages to criminal justice data should capture the cases in which people with FASD feature as victims, as well as perpetrators, to not give a distorted picture of their lives.

Clinicians and researchers highlighted there would be value in including data from cases where patients had been assessed for FASD but did not meet criteria, suggesting this could provide an important comparison group:

> *‘there’s something about capturing those who would deemed sufficiently at risk to warrant an assessment but ultimately it was deemed not to be’. (Clinician, P3, main workshop)*

> *‘I think the exposed group that don’t meet criteria is a really important comparator’ (Clinician, P17, main workshop)*

Additionally, clinicians described the importance of retaining flexibility and space for clinical judgement in their practice. They discussed the complexity of FASD diagnosis and expressed concerns that a solely quantitative dataset within a national FASD database could lead to the loss of important details from qualitative notes:

> *‘It would help that if there was some qualitative aspect or element [to the database]… a lot of the really juicy data that we get comes from the developmental history [and] triangulating between questionnaires, clinical observation, direct assessment’ (Clinician, P4, breakout room)*

> *‘[When doing clinical testing you have] to look at the quality of what [those being assessed] do and how they do it as much as the numbers themselves, numbers don’t always tell the truth…sometimes you have to go with your gut’ (Clinician P9, main workshop)*

As described in the *challenges* section above, clinicians reported having limited time for data input. Therefore, efficient data input procedures and consideration of additional support for initial data input was deemed an important database development.

### Data availability

Seven clinical stakeholders completed our FASD data capture proforma, based on the Canadian National Database for FASD (template provided in Supplementary appendix 1) [33]. This exercise was designed to assess the format and availability of FASD data in UK clinics and its potential compatibility with international databases. Overall, there was a large degree of alignment in the data collected in clinics with the measures in the data capture proforma; additionally, clinicians mostly agreed that, if these measures were not currently being collected, they would consider collecting this in the future. Stakeholders expressed a willingness to share personal identifier, demographic and clinical data, subject to robust governance processes. More details about the availability of different types of clinical data are presented below.

### Demographic information and patient characteristics

On average, the stakeholders who completed the data capture forms reported that they currently collect 64% of the demographic and patient characteristic measures listed on the proforma. For the measures where clinics did not collect data, on average, 75% stated that they would be willing to collect these data in the future, and 25% stated that they may consider collecting these data in the future. On average, stakeholders indicated that they would be willing to share 91% of the measures on the data capture proforma (subject to robust data governance), they were not willing to share 4% of the measures and 6% answered “maybe” when asked if they were willing to share. Measures which clinics were most reluctant to share included information on the source of referral, date of birth and living situation.

### Sibling and parent diagnosis of FASD

Forty nine percent of the stakeholders who completed the data capture forms collected data on sibling FASD diagnosis. Of those who didn’t collect this information, 80% answered that they would consider collecting data on sibling diagnoses and 20% answered that they would “maybe” consider collecting sibling data. Regarding sharing data on sibling diagnoses, 67% of clinicians were happy to share and 33% answered “maybe” when asked if they were willing to share these data. Thirty-three percent of clinicians who completed the proforma said that they collected data parent FASD diagnoses. Of those who didn’t collect these data 67% answered “yes” and 33% answered “maybe” when asked if they would consider collecting it in the future. Regarding sharing data on parental FASD diagnosis, 75% of clinicians were happy to share and 25% answered “maybe”.

### Sentinel facial features

Finally, regarding sentinel facial features, 100% of clinicians who completed the data capture proforma collected data on this and 100% were willing to share these data.

## DISCUSSION

Our multidisciplinary, multi-method public and professional involvement activities demonstrated broad support for the development of a pseudonymised national linked database for FASD in the UK. For people living with FASD and third-sector representatives, the main anticipated benefit was increased understanding, awareness, and support for FASD. Lived experience stakeholders expressed an urgent need for more data and research and communicated that a national FASD database could play an important part in achieving thisClinicians explained that a national database for FASD could provide important new insights into clinical profiles and that this had the potential to support more informed and efficient diagnosis. Researchers suggested that a linked FASD database could provide important novel insights into health, criminal justice and social outcomes associated with FASD over time, including the potential to identify opportunities for interventions to improve these outcomes. Policy-makers reported that the establishment of a national database for FASD in the UK would align well with current priorities including FASD and data transformation policies. All contributors expressed the importance of robust data governance and consent processes.

Key perceived challenges were the resource implications of harmonising, collating, and inputting existing data, which were described as highly variable in terms of availability and format. Clinicians were supportive of an element of standardisation for prospective data collection but expressed the importance of retaining space for clinical judgement and qualitative data alongside the potential insights gained from quantitative data analysis. Results from our clinical data capture proformas indicated that most of the clinics that responded captured the data necessary to establish a national linked database for FASD, including data on prenatal alcohol exposure, growth, neurodevelopmental outcomes, sentinel facial features and personal identifiers. Clinicians who did not currently collect these data indicated that they would be willing to collect and share such data in the future, subject to robust governance.

Although the UK has seen a recent influx of policy and guidance on FASD in the UK [8, 11, 14, 23, 24], the data necessary to fully understand the impact of, and plan effective service provision for FASD, is lacking. FASD databases have been established in the United States, Canada and Australia. These have been instrumental in understanding the characteristics and needs of those living with FASD in their populations, and for informing policy and practice [32–34, 36, 37]. Accordingly, these offer useful exemplars for the establishment of a UK equivalent, and for illustrating the impact that this may achieve. Our public and professional involvement work demonstrated that UK clinicians were broadly in favour of data standardisation, following a data capture method based on the proforma used by the Canadian National Database for FASD [33]. Furthermore, a recent international survey indicated that 91% of clinicians were in favour of adopting a unified approach for FASD assessment [48]. This offers further potential for future international collaborations and comparisons, if a FASD database were to be established in the UK.

Current international examples of FASD databases have not been linked to routinely collected health, social care and administrative data sources, instead relying on retrospective reporting and follow up surveys to assess outcomes among those living with FASD. Given the UK has a rich infrastructure of routine data[38], the development of a national linked database has the potential to provide world-leading insights into an array of long-term health, social care, economic, education and criminal justice outcomes associated with FASD. This could transform (inter)national policy, prevention, and service provision for FASD.

Public involvement and engagement have been recognised as an essential step in gaining public support in big data research, and for ensuring that such endeavours reflect public views and priorities [49–53]. Following best practice guidelines, we adopted a flexible, multimethod approach to make our public involvement work as inclusive and accessible as possible for different stakeholder groups, and involved contributors at the earliest stages of our project [44, 52]. However, we also experienced some challenges. Some adults with FASD and their supporters appeared to find the concept of an pseudonymised national database for FASD difficult to grasp, with some mistakenly thinking that their data would go directly to their GP, for example. Making the concept of a national linked database clearer will be something which we will take forward with future work, for example co-developing, with lived experience stakeholders, a clear and concise way of communicating this concept.

Similarly, Teodorowski and colleagues (2023) highlighted that big data can be an abstract and complex topic to discuss with the public, especially among ‘seldom-heard’ groups, including those with disabilities [49]. This issue is likely to be compounded in the case of FASD, as individuals with FASD often face challenges with abstract reasoning [12]. The literature suggests that animations and visualisations may make discussions around big data more accessible for the public [49]. This is something that we plan to use to a greater extent in future PPI work on this topic.

Given the varied responses of each group flexible, diverse, and embedded involvement of a range of stakeholders will be essential as we seek to develop a national database for FASD in the UK. Consistent with existing literature on public views on the use and linkage of patient data [54], our stakeholders gave mixed opinions about preferred consent models. While clinicians, data governance, academic and most individuals living with FASD expressed that a default ‘opt-out’ model for this pseudonymised database would be acceptable, some expressed concerns about their data being included without their explicit consent. Given the spectrum of opinion on preferred models of consent among stakeholders, ongoing stakeholder engagement will be essential as we seek to design a consent model that effectively balances the need to ensure a sustainable resource, that incurs minimal participant burden and which provides transparency in the process of consent, security, and use of relevant data.

The work presented in this paper has many strengths. We adopted a multi-disciplinary approach gaining perspectives from a range of stakeholders including adults with FASD, caregivers of people with FASD, clinicians, representatives from the third sector, data specialists, academics, and policy stakeholders. Each group offered different perspectives and ideas on the benefits, challenges and preferred design of the database. It was strongly felt among the project team that we would have missed important points if we had not captured data/views from all these groups. In order to reach these groups, we used a multi-method approach, encompassing a hybrid workshop, online 1:1 and group meetings, and email conversations. This multi-format approach enabled more contributors to join and reduced the potential burden of travel/time commitments, which may have been barriers to inclusion for some. Additionally, there was a range of skill sets within the project team, including researchers’ data expertise, quantitative and qualitative expertise, specialist clinical, and lived-experience insight. This enabled a broad understanding of the barriers and facilitators that the stakeholders presented to us.

A potential limitation of this work is that the results present high-level, descriptive summaries. As this was a PPI project and not a qualitative research study, this was an appropriate format to present the views and recommendations of our contributors, however we may have missed some of the insights that might have been gained from more in-depth qualitative analysis. Another potential limitation is that our workshops may have suffered from contagion and dominant voices may have led the conversation and others may not have felt they could share their views freely.

## CONCLUSIONS

Our multi-method and multidisciplinary public and professional involvement activities demonstrated broad support for the development of a pseudonymised national linked database for FASD in the UK. Our stakeholders reported that a national FASD database could have far-reaching benefits including facilitating advances in research and policy, improving prevention, and increasing awareness, understanding and effective support for FASD. Importantly this resource, would provide a step-change in increasing the accessibility and visibility of FASD in key public health data sources. Our contributors also highlighted some challenges mainly regarding the practicalities of using the database and data governance issues, and made recommendations for important design features. The perceived benefits and challenges of the database varied by stakeholder group demonstrating that flexible, diverse, embedded stakeholder collaboration will be essential as we seek to establish this database. Given the its relatively sophisticated routine data infrastructure, the UK has the potential to develop a world-leading resource to support advancement of FASD knowledge, policy and service provision.

## Data Availability

All data produced in the present study are available upon reasonable request to the authors

## Acknowledgments

The authors would like to thank the members of the public and professionals who provided us with their valuable knowledge and opinions. This work was funded by Jean Golding Institute (JGI) Seed Corn Funding (awarded to CM for PPI activities with professional stakeholders), Elizabeth Blackwell PPI funding through the Wellcome Trust Grant 204813/Z/16/Z (awarded to CM for lived experience workshops) and for National Institute for Health Research Biomedical Research Centre in Bristol (for project team data expertise funding and conference attendance) this funded SH and BS’s to do this work and SH was additionally supported by National Institute for Health Research Applied Research Collaboration West (NIHR ARC West). During this work, CM was supported by the National Institute for Health and Care Research (NIHR) School for Public Health Research (SPHR) (Grant Reference Number PD-SPH-2015). AB was additionally supported by the Health Data Research UK (Ref: HDRUK2023) which is the UK’s Health Data Research institute (which is funded by UK Research and Innovation, the Medical Research Council, the British Heart Foundation, Cancer Research UK, the National Institute for Health and Care Research, the Economic and Social Research Council, the Engineering and Physical Sciences Research Council, Health and Care Research Wales, Health and Social Care Research and Development Division [Public Health Agency, Northern Ireland], Chief Scientist Office of the Scottish Government Health and Social Care Directorate). SB was employed by The National Organisation for FASD and supported by the Sylvia Adams Charitable Trust, Four Acres Trust, Contact/Pears, Diageo and other supporters. PC was supported by the NIHR and the Oglesby Charitable Trust and the University of Salford. RM was primarily funded by the NHS but also receives funding from NIHR and Ogelsby Trust for FASD. The views expressed are those of the author(s) and not necessarily those of any of the funders mentioned above. We are all very grateful for the support and funding received.

## Ethics statement

These activities were classed as public involvement work, rather than research. In this work, members of the public were acting as specialist advisors, providing valuable knowledge and expertise based on their experience of a health condition or public health concern. Public contributors received written details of the planned activities and were given the opportunities to discuss their involvement with the project team before they participated in each session. However, review by a research ethics committee and formal processes for obtaining informed consent were not needed (NIRH involve – <u>Do I need to apply for ethical</u> <u>approval to involve the public in my research? | INVOLVE</u>).

## Data access statement

Data is not available as this paper is a write up of PPI insights and not data.

## Abbreviations

DHSC: Department of Health and Social Care
FAS: Fetal alcohol syndrome
FASD: fetal alcohol spectrum disorder
NHS: National Health Service
NICE: National Institute for Health and Care Excellence
OHID: Office for Health Improvement and Disparities
SNOMED CT: Systematized Nomenclature of Medicine Clinical Terms
UK: United Kingdom
UKSPI: UK Standards for Public Involvement in Research

## References

1. Popova S, Charness ME, Burd L, Crawford A, Hoyme HE, Mukherjee RAS, et al. Fetal alcohol spectrum disorders. Nature Reviews Disease Primers. 2023;9(1):11.

2. McCarthy R, Mukherjee RA, Fleming KM, Green J, Clayton-Smith J, Price AD, et al. Prevalence of fetal alcohol spectrum disorder in Greater Manchester, UK: An active case ascertainment study. Alcoholism: Clinical and Experimental Research. 2021;45(11):2271–81.

3. Lange S, Probst C, Gmel G, Rehm J, Burd L, Popova S. Global Prevalence of Fetal Alcohol Spectrum Disorder Among Children and Youth: A Systematic Review and Meta-analysis. JAMA Pediatrics. 2017;171(10):948–56.

4. Streissguth AP, Bookstein FL, Barr HM, Sampson PD, O’Malley K, Young JK. Risk factors for adverse life outcomes in fetal alcohol syndrome and fetal alcohol effects. Journal of Developmental & Behavioral Pediatrics. 2004;25(4):228–38.

5. Easton B, Burd L, Rehm J, Popova S. Productivity losses associated with Fetal Alcohol Spectrum Disorder in New Zealand. N Z Med J. 2016;129(1440):72–83.

6. Greenmyer JR, Klug MG, Kambeitz C, Popova S, Burd L. A Multicountry Updated Assessment of the Economic Impact of Fetal Alcohol Spectrum Disorder: Costs for Children and Adults. Journal of Addiction Medicine. 2018;12(6).

7. Popova S, Lange S, Shield K, Mihic A, Chudley AE, Mukherjee RAS, et al. Comorbidity of fetal alcohol spectrum disorder: a systematic review and meta-analysis. Lancet. 2016;387(10022):978–87.

8. Scottish Intercollegiate Guidelines Network (SIGN). Children and young people exposed prenatally to alcohol (SIGN 156): a national clinical guideline. 2019.

9. Popova S, Dozet D, Burd L. Fetal Alcohol Spectrum Disorder: Can We Change the Future? Alcoholism: Clinical and Experimental Research. 2020;44(4):815–9.

10. Scholin L, Mukherjee RAS, Aiton N, Blackburn C, Brown S, Flemming KM, et al. Fetal alcohol spectrum disorders: an overview of current evidence and activities in the UK. Archives of disease in childhood. 2021;106(7):636–40.

11. National Institute for Health and Care Excellence. Fetal alcohol spectrum disorder. Quality standard [QS204]. 2022.

12. British Medical Association. Alcohol in pregnancy: preventing and managing fetal alcohol spectrum disorders. 2016.

13. NHS England. SNOMED CT 2023 [Available from: https://digital.nhs.uk/services/terminology-and-classifications/snomed-ct.

14. Department of Health and Social Care. Fetal alcohol spectrum disorder: health needs assessment. 2021.

15. NHS Digital. SNOMED Code Usage in Primary Care, 2021-22 2022 [Available from: https://digital.nhs.uk/data-and-information/publications/statistical/mi-snomed-code-usage-in-primary-care/2021-22.

16. World Health Organisation. International Classification of Diseases, Eleventh Revision (ICD-11) 2019 [

17. May PA, Gossage JP, Kalberg WO, Robinson LK, Buckley D, Manning M, Hoyme HE. Prevalence and epidemiologic characteristics of FASD from various research methods with an emphasis on recent in-school studies. Developmental disabilities research reviews. 2009;15(3):176–92.

18. Morleo M, Woolfall K, Dedman D, Mukherjee R, Bellis MA, Cook PA. Under-reporting of foetal alcohol spectrum disorders: an analysis of hospital episode statistics. BMC Pediatr. 2011;11:14.

19. Steer C, Campbell, H., Cockburn, F., McClure, J., McIntosh, N.,, editor The incidence of fetal alcohol syndrome in Scotland: an enhanced passive surveillance study. Fourth European Conference on FASD; 2016; Royal Holloway, University of London.

20. Mukherjee R, Wray E, Curfs L, Hollins S. Knowledge and opinions of professional groups concerning FASD in the UK. Adoption & Fostering. 2015;39(3):212–24.

21. Young S, Absoud M, Blackburn C, Branney P, Colley B, Farrag E, et al. Guidelines for identification and treatment of individuals with attention deficit/hyperactivity disorder and associated fetal alcohol spectrum disorders based upon expert consensus. BMC Psychiatry. 2016;16(1):324.

22. Burleigh CR, Lynn RM, Verity C, Winstone AM, White SR, Johnson K. Fetal alcohol syndrome in the UK. Arch Dis Child. 2023;108(10):852–6.

23. National Organisation for FASD. The time is now: the national perspective on ramping up FASD prevention, diagnosis and support services. 2022.

24. Public Health England (PHE). Maternity high impact area: Reducing the incidence of harms caused by alcohol in pregnancy. 2020.

25. Department of Health and Social Care. Data saves lives: reshaping health and social care with data. 2022.

26. Department of Health and Social Care. A plan for digital health and social care. 2022.

27. Central Digital and Data Office. Transforming for a digital future: 2022 to 2025 roadmap for digital and data - updated September 2023.; 2023.

28. Government Analysis Function. Reproducible Analytical Pipelines (RAP) strategy. 2022.

29. NHS England. NHS Federated Data Platform 2022 [Available from: https://www.find-tender.service.gov.uk/Notice/008755-2022?origin=SearchResults&p=1.

30. Welsh Government. Digital and data strategy for health and social care in Wales. Welsh Government; 2023.

31. Health and Social Care (HSC) Northern Ireland. Digital Strategy Health and Social Care Northern Ireland 2022-2030 2022 [Available from: https://www.health-ni.gov.uk/publications/digital-strategy-health-and-social-care-northern-ireland-2022-2030.

32. Cook J, Unsworth K, Flannigan K. Characterising fetal alcohol spectrum disorder in Canada: a national database protocol study. BMJ Open. 2021;11(9):e046071.

33. Canada Fetal Alcohol Spectrum Disorder Research Network. 2018. [October 12, 2023]. Available from: https://canfasd.ca/2018/07/18/the-national-fasd-database/.

34. The Australian Paediatric Surveillance Unit. Fetal Alcohol Spectrum Disorder Australian Registry (FASDAR) 2023 [Available from: https://www.apsu.org.au/home/fetal-alcohol-spectrum-disorder-australian-registry-/.

35. University of Alaska Anchorage College of Health. Alaska Fetal Alcohol Spectrum Disorders Data Systems Development: Gaps, Opportunities, & Recommendations. 2021.

36. Astley SJ. Profile of the first 1,400 patients receiving diagnostic evaluations for fetal alcohol spectrum disorder at the Washington State Fetal Alcohol Syndrome Diagnostic & Prevention Network. Canadian Journal of Clinical Pharmacology. 2010;17(1):e132–64.

37. Astley SJ, Bailey D, Talbot C, Clarren SK. Fetal alcohol syndrome (FAS) primary prevention through fas diagnosis: II. A comprehensive profile of 80 birth mothers of children with FAS. Alcohol Alcohol. 2000;35(5):509–19.

38. Health Data Research UK (HDR UK). The Sudlow Review: ’Unifying Health Data in the UK’ HDR UK; 2023 [Available from: https://www.hdruk.ac.uk/helping-with-health-data/the-sudlow-review/.

39. Jonsson E, Salmon A, Warren KR. The international charter on prevention of fetal alcohol spectrum disorder. The Lancet Global Health. 2014;2(3):135–7.

40. Carter P, Laurie GT, Dixon-Woods M. The social licence for research: why care. data ran into trouble. Journal of medical ethics. 2015;41(5):404–9.

41. Teodorowski P, Jones E, Tahir N, Ahmed S, Frith L. Public involvement and engagement in big data research: protocol for a scoping review and a systematic review of delivery and effectiveness of strategies for involvement and engagement. BMJ open. 2021;11(8):e050167.

42. Waind E. Trust, security and public interest: striking the balance A narrative review of previous literature on public attitudes towards the sharing, linking and use of administrative data for research. International journal of population data science. 2020;5(3):1368.

43. National Institute for Health and Care Research. Briefing notes for researchers - public involvement in NHS, health and social care research 2021 [Available from: https://www.nihr.ac.uk/documents/briefing-notes-for-researchers-public-involvement-in-nhs-health-and-social-care-research/27371.

44. UK Public Involvement Standards Development Partnership. UK Standards for Public Involvement in Research 2019 [Available from: https://sites.google.com/nihr.ac.uk/pi-standards/standards.

45. Staniszewska S, Brett J, Simera I, Seers K, Mockford C, Goodlad S, et al. GRIPP2 reporting checklists: tools to improve reporting of patient and public involvement in research. BMJ. 2017;358:j3453.

46. NHS England and NHS Improvement. Quality, Service Improvement and Redesign Tools: Stakeholder analysis.

47. National Institute for Health and Care Research. Payment guidance for researchers and professionals. 2022.

48. Reid N, Shanley DC, Logan J, White C, Liu W, Hawkins E. International Survey of Specialist Fetal Alcohol Spectrum Disorder Diagnostic Clinics: Comparison of Diagnostic Approach and Considerations Regarding the Potential for Unification. Int J Environ Res Public Health. 2022;19(23).

49. Teodorowski P, Rodgers SE, Fleming K, Tahir N, Ahmed S, Frith L. ’To me, it’s ones and zeros, but in reality that one is death’: A qualitative study exploring researchers’ experience of involving and engaging seldom-heard communities in big data research. Health expectations : an international journal of public participation in health care and health policy. 2023;26(2):882–91.

50. Jones KH, Heys S, Thompson R, Cross L, Ford D. Public involvement & engagement in the work of a data safe haven: a case study of the SAIL Databank. International journal of population data science. 2020;5(3):1371.

51. Finlay-Jones A, Symons M, Tsang W, Mullan R, Jones H, McKenzie A, et al. Community priority setting for fetal alcohol spectrum disorder research in Australia. International Journal of Population Data Science. 2020;5(3).

52. Aitken M, Tully MP, Porteous C, Denegri S, Cunningham-Burley S, Banner N, et al. Consensus Statement on Public Involvement and Engagement with Data Intensive Health Research. International journal of population data science. 2019;4(1):586.

53. Aitken M, McAteer G, Davidson S, Frostick C, Cunningham-Burley S. Public Preferences regarding Data Linkage for Health Research: A Discrete Choice Experiment. International journal of population data science. 2018;3(1):429.

54. ARDUK. Trust, Security and Public Interest: Striking the Balance. A review of previous literature on public attitudes towards the sharing and linking of administrative data for research. 2020.

